# Assessment of Machine Learning Algorithms for Prediction of Breast Cancer Malignancy Based on Mammogram Numeric Data

**DOI:** 10.1101/2020.01.08.20016949

**Authors:** Peter T. Habib, Alsamman M. Alsamman, Sameh E. Hassnein, Ghada A. Shereif, Aladdin Hamwieh

## Abstract

in 2019, estimated New Cases 268.600, Breast cancer has one of the most common cancers and is one of the world’s leading causes of death for women. Classification and data mining is an efficient way to classify information. Particularly in the medical field where prediction techniques are commonly used for early detection and effective treatment in diagnosis and research.These paper tests models for the mammogram analysis of breast cancer information from 23 of the more widely used machine learning algorithms such as Decision Tree, Random forest, K-nearest neighbors and support vector machine. The spontaneously splits results are distributed from a replicated 10-fold cross-validation method. The accuracy calculated by Regression Metrics such as Mean Absolute Error, Mean Squared Error, R2 Score and Clustering Metrics such as Adjusted Rand Index, Homogeneity, V-measure.accuracy has been checked F-Measure, AUC, and Cross-Validation. Thus, proper identification of patients with breast cancer would create care opportunities, for example, the supervision and the implementation of intervention plans could benefit the quality of long-term care. Experimental results reveal that the maximum precision 100%with the lowest error rate is obtained with Ada-boost Classifier.

## 1. Introduction

More people are dying annually from non-communicable diseases than from infectious diseases at this time. Breast cancer is the prominent and leading cause of women’s death worldwide among non-communicable diseases. In 2019,6.9%of the cancer deaths were due to breast cancer in which approximately 41,760 people died this year only(https://seer.cancer.gov/statfacts/html/breast.html).

The risk of breast cancer lies in every woman. When she lives at 85, one in eight(12%)is likely to develop breast cancer sometime in her lifetime. When a woman ages, she increases dramatically her risk of developing breast cancer independently of her family history[1].Most of the cancer development caused by mutation, especially single nucleotide polymorphism(SNP), in genes such as BRCA1which is the most common gene occurs in breast and ovarian cancer[2]

The aim of early diagnostic strategies is not only to provide quick access to cancer treatment but also to promote reliable diagnostic facilities. Screening requires the detection of tumors through screening people for signs before they are affected. For many decades mammography has been the gold standard form for testing over breast cancer diagnosis and mortality reduction[3].

Clinical outcome predictions based on machine learning may be used to decide properly and may result in better care for patients. Prediction models of machine learning can reliably identify people who should undergo biopsy and help reduce missing women who will die from breast cancer.ML also has a huge advantage over traditional statistical models, such as high power and disease prediction reliability. To our understanding, the prediction model does not have a particular algorithm that does better. We thus have conducted the most popular algorithms in the prediction of breast cancer and have contrasted their performance[4].

In this study, the breast cancer dataset is applied to 23 machine learning algorithms to identify as benign and malignant. To measure the performance of the algorithms we used the Wisconsin breast cancer dataset from the UCI Machine Learning Repository[5]. There is a suggestion that the algorithm provided an algorithm of the accuracy of 98,24%,99,63%,100%, for 50-50,60-40 and 70-30 respectively, and accuracy of classification of 100%for the 10 cross-validation scheme. Measures such as F1 Score which is needed for seeking a balance between Precision and Recall, AUC curves summarize the trade-off between the true positive rate and false-positive rate for a predictive model using different probability thresholds, Area under the curves(AUC)are used to validate the performance. The results show that Ada-boost Classifier works well with the breast cancer database and can be a good alternative to the well-known machine learning methods.

## 2. Materials and Methods

### 2.1 Collecting data

Data from the UCI learning machine database were collected. For the intent of this study, secure personal health information has been deleted. This study was exempt from the ethics review by the Ethics Committee of CHUC since it analyzed de-identified data, and all participants agreed in writing before the study was entered.

### 2.2 Breast cancer dataset

The Wisconsin Breast Cancer(original)datasets 20 from the UCI Machine Learning Repository is used in this study.Breast cancer Wisconsin has 570 instances(Benign:357 Malignant:212),2 classes(37.19%malignant and 62.63%benign),and 32 integer-valued attributes.

### 2.3 Experiment Framework

All classifier experiments mentioned in this paper are carried out with Scikit-learn libraries. (formerly scikits.learn and also known as sklearn).Scikit-learn comprises a set of machine learning algorithms to pre-process, detect, regressive, cluster and associate rules on results. Several real-world problems are discussed through Machine Learning methods in Scikit-learn. The paper provides experimenters and developers with a well-defined framework to construct and assess their models.

## 3. Results

### 3.1 Effectiveness

To apply and validate our classifiers, we employ a 10-fold cross-validation method, a methodology used to analyze predictive models, that broke the initial into a sample for model testing and training data for model evaluation. Upon implementing the pre-processing and planning methods we attempt to visually analyze the data to assess the quality and performance distribution of the values.Ada-boost Classifyers accuracy testing shows that 100%,97.4%,95.9%and 94.7%accuracy on 10%,20%,40%and 50%respectively.and other algorithms are shown in the following figure(1)

**Figure1.**
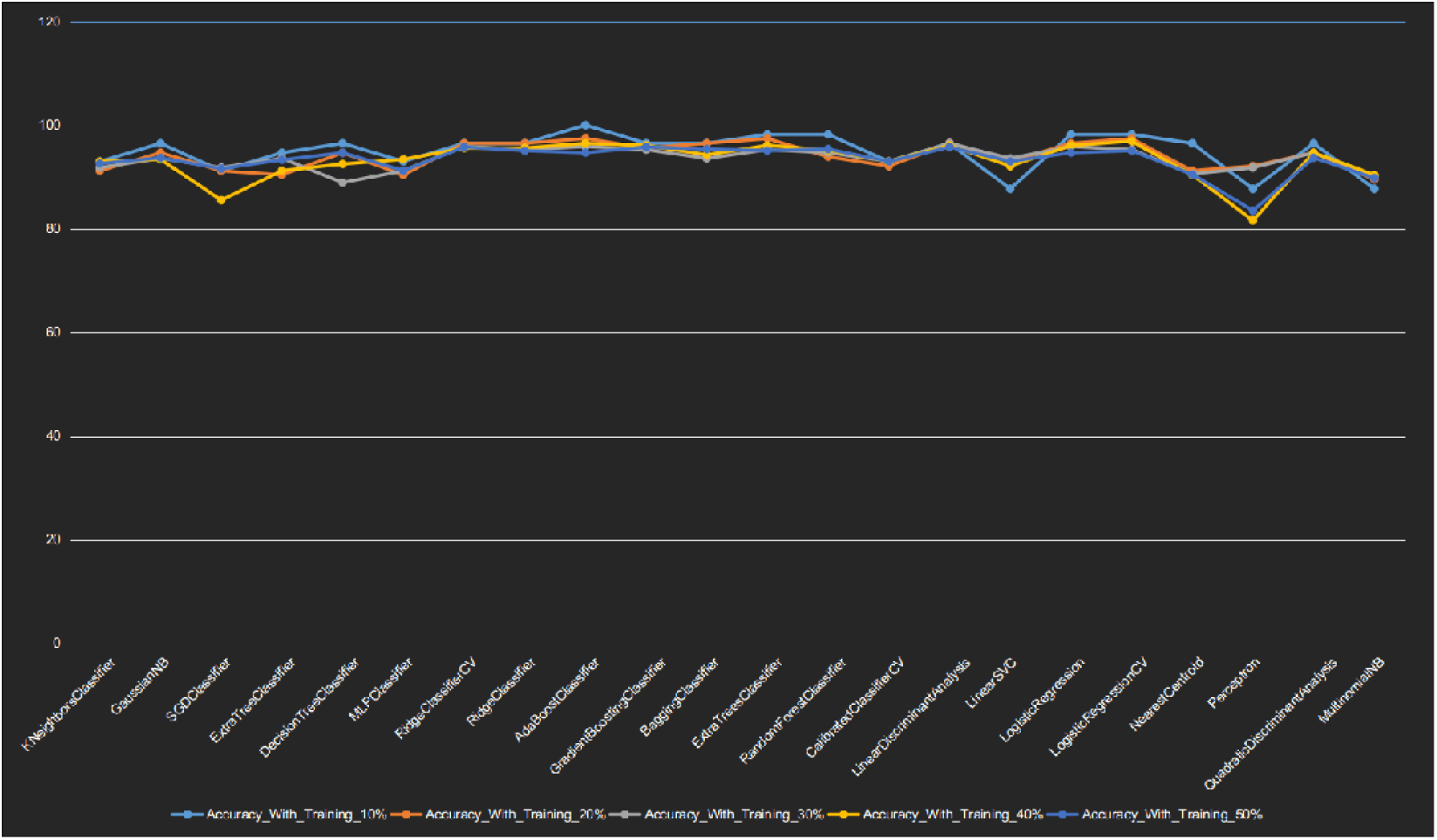

Simulation failure is also regarded in this study to help assess the quality of classifiers. To do so, we evaluate the effectiveness of our classifier in terms of:

1. Mean Absolute Error,
2. Mean Squared Error,
3. R^2^ Score,
4. Homogeneity Score
5. Adjusted Rand Score
6. V-Measure Score

The results are shown in Fig.2.After analysis of produced data,Ada-boost Classifier shows 100%,100%,100%,100%,0,and 0 with Adjusted Rand Score,Homogeneity Score,R^2^ Score,V-Measure,Mean Absolute Error,and Mean Squared Error respectively.

**Figure 2:**
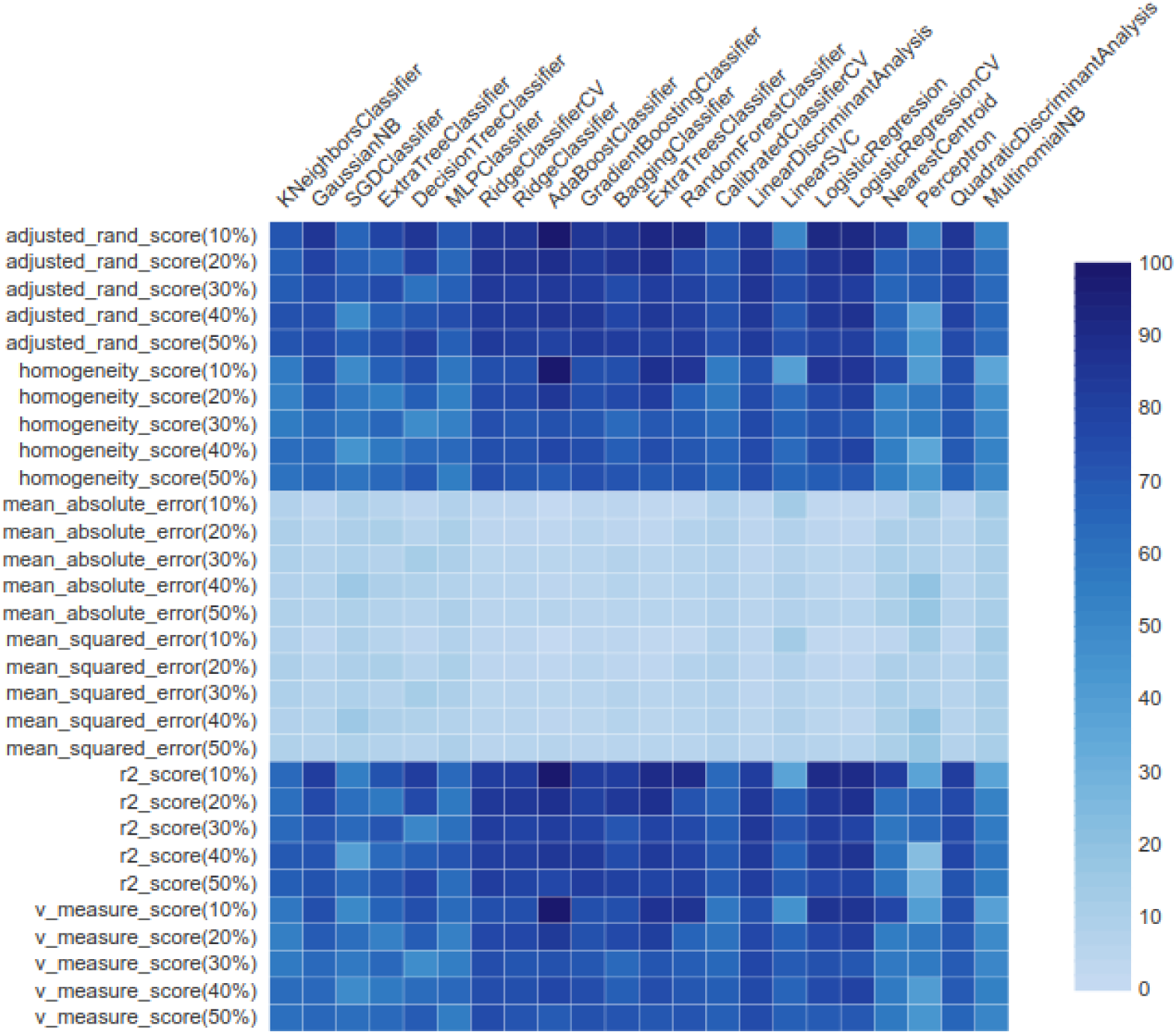
Heat-map shows score of each algorithm with different matrices.

### 3.2 Efficiency

Once the predictive model is done, we can test how functional it is. For that, we compare the accuracy measures based on AUC and F-Measure rate values for the algorithm as shown in Table(2) and figure(3). Ada-boost Classifier shows the best numbers comparing with other algorithms.

**Table1:** calculated accuracy scores according to different matrices

| Matrices used with different training set | Estimated Percentages(%) |
| --- | --- |
| adjusted_rand_score(10%) | 100 |
| adjusted_rand_score(20%) | 89.6 |
| adjusted_rand_score(30%) | 84.1 |
| adjusted_rand_score(40%) | 86.4 |
| adjusted_rand_score(50%) | 79.9 |
| homogeneity_score(10%) | 100 |
| homogeneity_score(20%) | 85.2 |
| homogeneity_score(30%) | 73.6 |
| homogeneity_score(40%) | 77.8 |
| homogeneity_score(50%) | 69.5 |
| mean_absolute_error(10%) | 0 |
| mean_absolute_error(20%) | 2.6 |
| mean_absolute_error(30%) | 4.1 |
| mean_absolute_error(40%) | 3.5 |
| mean_absolute_error(50%) | 5.3 |
| mean_squared_error(10%) | 0 |
| mean_squared_error(20%) | 2.6 |
| mean_squared_error(30%) | 4.1 |
| mean_squared_error(40%) | 3.5 |
| mean_squared_error(50%) | 5.3 |
| r2_score(10%) | 100 |
| r2_score(20%) | 88.3 |
| r2_score(30%) | 81.9 |
| r2_score(40%) | 85.2 |
| r2_score(50%) | 77.4 |
| v_measure_score(10%) | 100 |
| v_measure_score(20%) | 84.2 |
| v_measure_score(30%) | 73.8 |
| v_measure_score(40%) | 77.6 |
| v_measure_score(50%) | 69.2 |

**Table2:** Estimated values for Ada-boost Classifier presented in percent with time presented in seconds

| Ada-boost Classifier with<br>different training sets | Estimated<br>Values |
| --- | --- |
| AUC(10%) | 100 |
| AUC(20%) | 96 |
| AUC(30%) | 94 |
| AUC(40%) | 96 |
| AUC(50%) | 93 |
| F-Measure(10%) | 100 |
| F-Measure(20%) | 98 |
| F-Measure(30%) | 95 |
| F-Measure(40%) | 97 |
| F-Measure(50%) | 95 |
| No Skill | 50 |
| TimeToBuild(10%) | 0.16 |
| TimeToBuild(20%) | 0.15 |
| TimeToBuild(30%) | 0.14 |
| TimeToBuild(40%) | 0.15 |
| TimeToBuild(50%) | 0.12 |

**Figure3.**
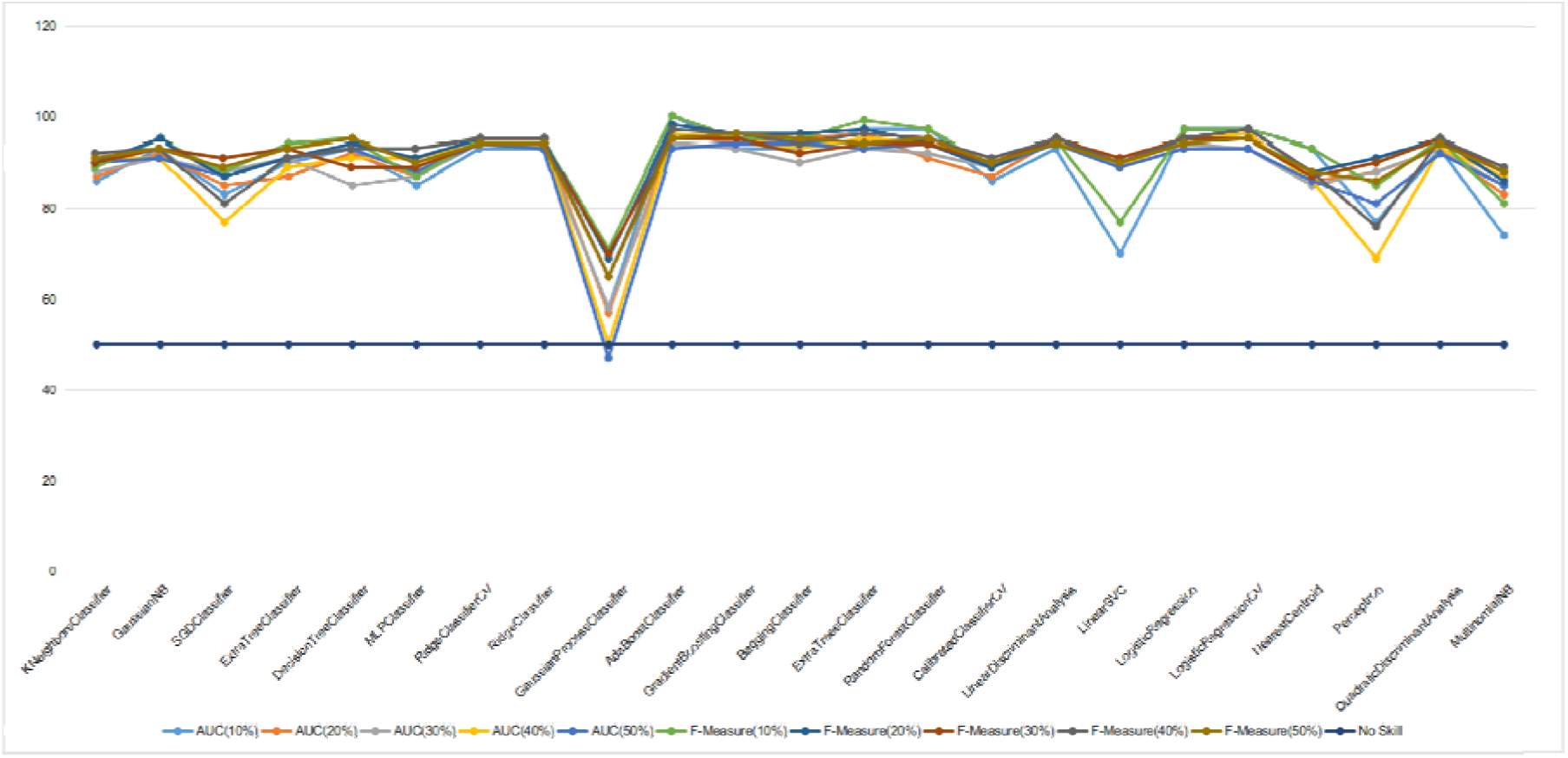

## 4. Discussion

Ensemble algorithm is a composite model that combines many low classification approaches to create the best classification system. Ensemble provides greater accuracy than single or generic classifiers. Procedures will coordinate by assigning each base learner to different machines. in the end, the ensemble of training methods is meta-algorithms, integrating many methods of machine learning in a common predictive model to improve efficiency. Combined approaches may decrease the uncertainty by bagging, bias by boosting or enhance predictions by stacking The following is how Ada-boost Classification works:

1. Ada-boost first randomly selects a section of training data for learning.
2. It exercises the Ada-boost learning system in an iterative manner with the choice of a training set dependent on an exact forecast for the last testing.
3. This assigns the greatest weight for misclassified findings so that these results are highly likely to be identified in the next version.
4. In every iteration, it also assigns a weight to the best classifier according to the classifier’s accuracy. A more reliable weight is given to the classifier.
5. 5-This method is iterated until full training information suits without error or the defined peak estimators have been reached.
6. To identify, make a “poll” for all the research algorithms that you have made.

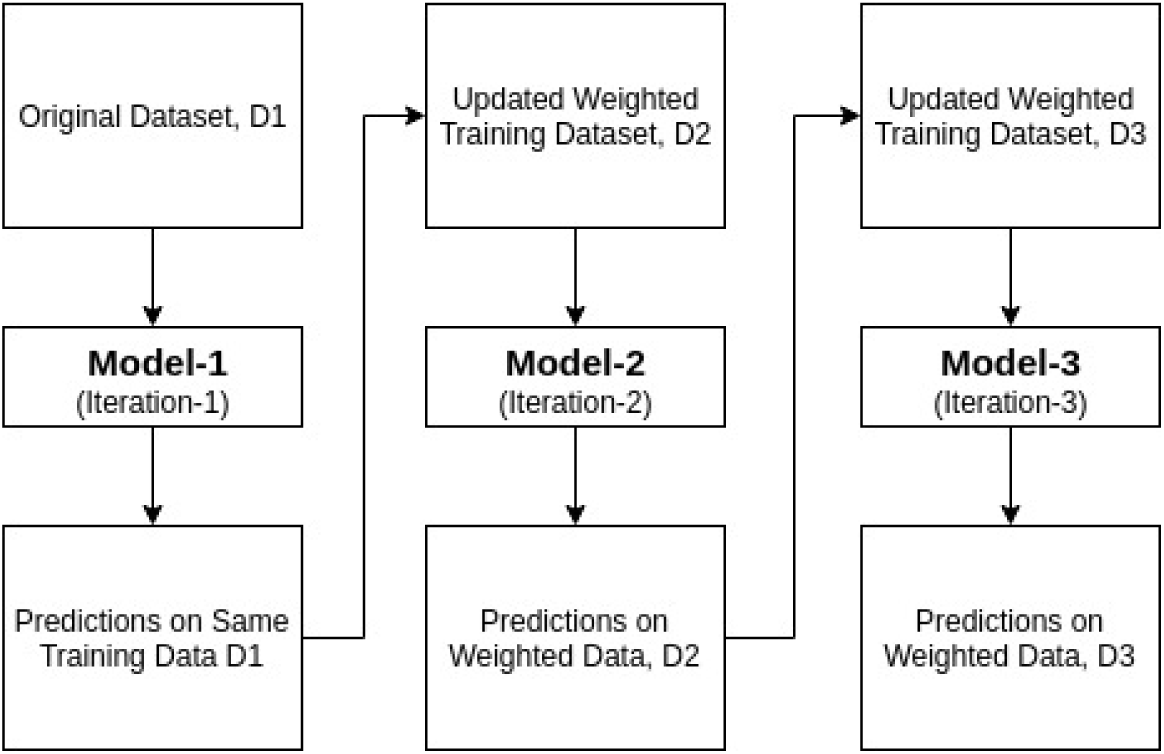

## 5. Conclusion

In 1996, Yoav Freund and Robert Schapire [7] proposed Ada-boost or Adaptive Boosting is one of the ensemble boosting classifier. This incorporation of multiple classifiers to boost classifiers performance. Ada-boost is a tool for the iterative collection. By combining several badly performing classifiers, the Ada-Boost Classifier builds a strong classifier to yield strong accuracy. Ada-boost’s basic concept is to set classifiers weights and to train the sample data in each step so that unexpected results can be predicted accurately. Any algorithm for machine learning can be used as a simple classifier if weight is acknowledged.

## Data Availability

https://github.com/peterhabib/EvaluationOfML/tree/master

## 6. Software availability

Source code available from GitHub: https://github.com/peterhabib/EvaluationOfML/tree/master Archived source code as at the time of publication: 10.5281/zenodo.3572077. (6). License: MIT

## 7. Data availability

*Underlying data*: https://archive.ics.uci.edu/ml/datasets/breast+cancer+wisconsin+(original)

### Extended data

Scripts and Data sets used in Study: Scripts and Data sets used in Study, DOI: 10.5281/zenodo.3572077., License: Creative Commons Attribution 4.0 International

This project contains the following extended data:

accurecy.csv

breastCancer.csv

Data.csv

Estmatorxlsx.csv

EvaluationFile.csv

EvaluationFile(Normalized).csv

EvaluationFile(Scaled).csv

Matrices.csv

## 8. Author contributions

Peter T. Habib responsible for Conceptualization, Methodology, and developed Script of study

Alsamman M. Alsamman and Peter t. Habib contribute to Writing – Original Draft Preparation

And Writing – Review & Editing

Ghada A. Shereif contribute to Validation

Aladdin Hamwieh contribute to Supervision and funding of this paper

---

**Figure4: Accuracy of algorithms calculated with algorithm scoring matrix**

**Figure5: illustration of calculated algorithms without plotting time to avoid disruption**.

